# Waning, Boosting and a Path to Endemicity for SARS-CoV-2

**DOI:** 10.1101/2021.11.05.21265977

**Authors:** Matt J. Keeling, Amy Thomas, Edward M. Hill, Robin N. Thompson, Louise Dyson, Michael J. Tildesley, Sam Moore

## Abstract

In many countries, an extensive vaccination programme has substantially reduced the public-health impact of SARS-CoV-2, limiting the number of hospital admissions and deaths compared to an unmitigated epidemic. Ensuring a low-risk transition from the current situation to one in which SARS-CoV-2 is endemic requires maintenance of high levels of population immunity. The observed waning of vaccine efficacy over time suggests that booster doses may be required to maintain population immunity especially in the most vulnerable groups. Here, using data and models for England, we consider the dynamics of COVID-19 over a two-year time-frame, and the role that booster vaccinations can play in mitigating the worst effects. We find that boosters are necessary to suppress the imminent wave of infections that would be generated by waning vaccine efficacy. Projecting further into the future, the optimal deployment of boosters is highly sensitive to their long-term action. If protection from boosters wanes slowly (akin to protection following infection) then a single booster dose to the over 50s may be all that is needed over the next two-years. However, if protection wanes more rapidly (akin to protection following second dose vaccination) then annual or even biannual boosters are required to limit subsequent epidemic peaks an reduce the pressure on public health services.

---

The SARS-CoV-2 pandemic has had a devastating effect on mortality, morbidity, social wellbeing and economics worldwide [1-6]. Given the scale of the pandemic, eradication (either nationally or globally) seems unlikely over medium time-scales. It is therefore important to consider the transition from pandemic to endemic and how this can be managed without allowing excessive amounts of disease or resorting to repeated lockdowns. Maintaining high levels of population immunity is key to a successful transition to endemicity, yet this is confounded by waning immunity [7-8] - both following infection and vaccination. Here, using England as a well-described example with high quality data, we use fitted epidemiological models to consider the interplay between waning, boosters and infection dynamics over the next two years.

England has experienced three relatively synchronised waves of infection. The first wave began in early March 2020, reaching a peak of 3099 hospital admissions on 1st April 2020 then declining to relatively low levels by the summer of 2020. The second wave began in September of 2020 and was exacerbated by the emergence of the Alpha variant [9-11]; it reached a peak of 4134 hospital admissions on 12th January 2021 before declining. The third wave, largely associated with the Delta variant [12-13], began in June 2021 and can be characterised by its persistence and relatively low levels of hospital admissions (compared with cases), attributable to the success of the national vaccination campaign. Vaccination in England began on 8th December 2020, with a prioritisation schedule based on age and risk [14-15], and has generally seen high levels of vaccine uptake. By October 2021, over 2.6 million second doses had been administered to those aged 80 and over (approximately 97% of the age-group^1^) and over 10 million second doses had been given to those aged 60-79 (approximately 99% of the age-group^1^). The uptake has been lower within younger age groups (approximate 83% for 20-59 year olds), but has still contributed to substantially increasing population level immunity, limiting cases, hospitalisations and deaths in the community [16].

Multiple immunological studies have demonstrated an armory of protective immune responses against SARS-CoV-2 (specific antibodies, CD4+ T-cells and CD8+ T-cells [17,18]): however, immunological and epidemiological data show that immunity is not permanent. Neutralising serum antibodies are considered as correlates of protection against infection [8], whilst levels of binding antibody have been shown to correlate reasonably well with functional, neutralising antibodies [18]. Accordingly, antibody decay rates have been used as proxies for waning immunity, with SARS-CoV-2 specific antibodies having a half-life of around 100 days [8,19-22] and reduced levels indicative of a reduction in protection [8]. From observational epidemiological studies, there is evidence of repeated infection [23,24], which may be exacerbated by differences between variants, such that those infected with the wild-type variant are less immune to the Delta variant and are at risk of reinfection. Assessment of vaccine efficacy over time also shows a decline in protection against infection, symptoms and severe disease [25,26]. This decline is most pronounced in the elderly and vulnerable who were amongst the first to be vaccinated in the UK and therefore have had longer for efficacy to wane [25,26]. However, this picture is confounded by the use of different vaccines which generate different initial levels of protection and may wane at different rates. To combat this declining efficacy, a booster campaign primarily targeted at those over 50 years old (or in specific clinically vulnerable groups) has been launched, with the aim of increasing immunity in those most likely to suffer severe disease.

Here, using a mathematical model that has been fitted to the SARS-CoV-2 outbreak in England, we consider the role of the Autumn 2021 booster campaign (which initially targets those over 50, healthcare workers and those that are clinically vulnerable, at least 6 months after their second vaccine dose), whether additional vaccination programmes will be needed and what form these should ideally take. The model is a deterministic age-structured model [27] that also incorporates multiple variants [28] and vaccination [29]. It has been extended to include waning of immunity (both following infection and following vaccination) and third (booster) doses of vaccine (full details given in the Supplementary Material). In projecting this model forwards, we face three major sources of uncertainty.

Firstly, the mixing and social behaviour of the population has been seen to vary through time as different control measures are enforced or recommended. However, any change in the population’s level of precautionary behaviour (which is a combination of multiple mitigation measures e.g. social distancing, mask use, avoiding high-risk settings, working from home) is not simply a reflection of the current rules, but is strongly influenced by perceived risk. Historic levels of precautionary behaviour are inferred from the pattern of cases, hospital admissions and deaths (Figure 1) but a number of future projections are feasible. Here we simply extrapolate the linear decline that has been inferred since the beginning of 2021, leading to pre-COVID mixing in the younger population by March 2022 (CI December 2021-May 2022) and in the older population by April 2022 (CI January - August 2022).

**Figure 1.**
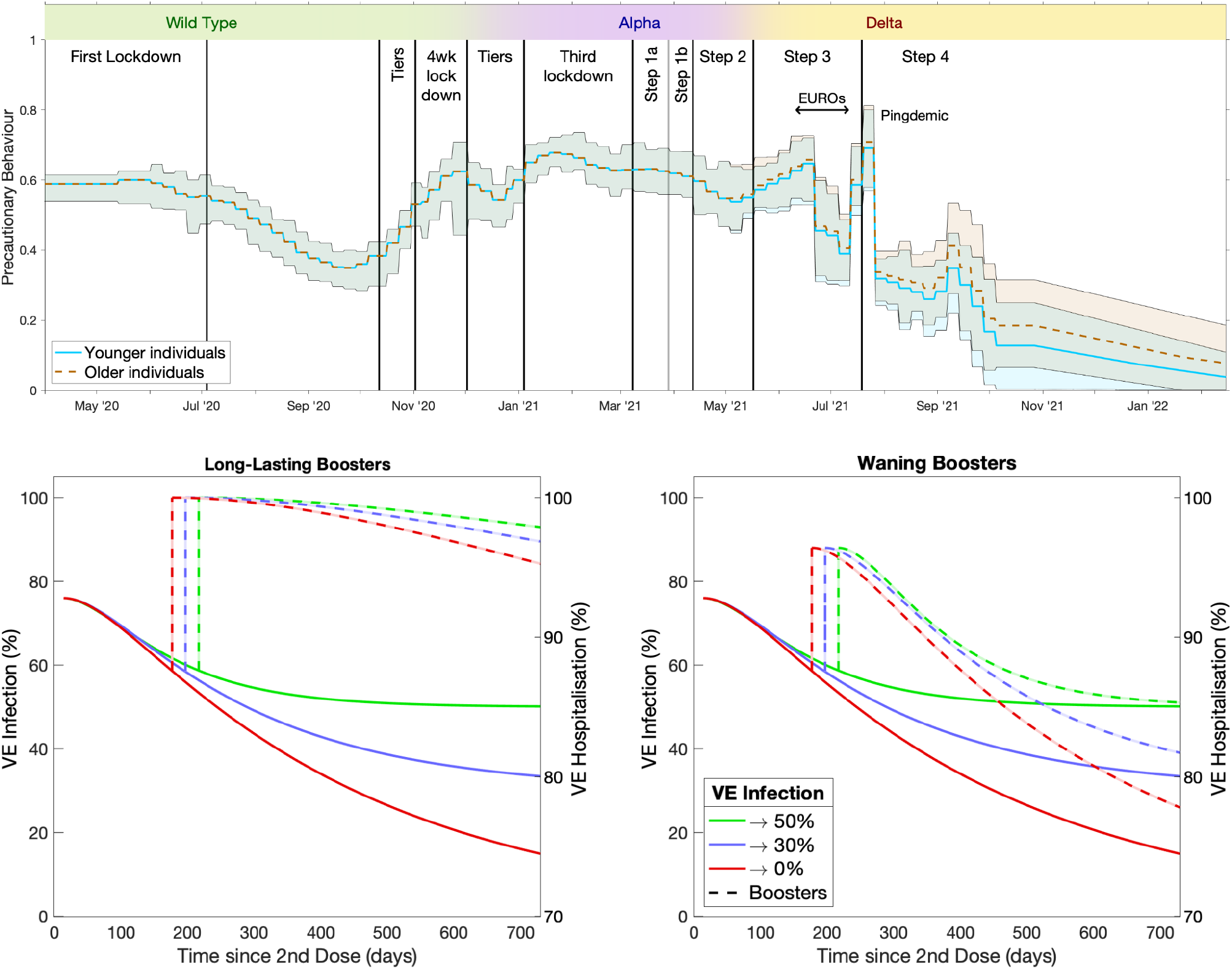
Inferred precautionary behaviour (top) and the range of vaccine efficacy assumptions (bottom). The inferred precautionary behaviour (blue solid line for younger people (<40 years old), dashed orange line for older individuals (>60 years old) together with 95% credible intervals) is shown together with the different periods of restrictions / behaviour and the three main variants in circulation (top panel, coloured bar). The lower figures show the assumed efficacy against infection, hospitalisation (solid lines: asymptoting to 50% (green), 30% (blue) and 0% (red) protection against infection) assuming equal amounts of Pfizer (BNT162b2) and AstraZeneca (ChAdOx1) vaccines and the action of boosters (dashed lines: high longer-lasting protection on the left; lower, waning protection on the right). Protection against death is assumed to wane in the same proportional way. The displayed timing of boosters is for illustrative purposes only.

Secondly, although data indicate that vaccine efficacy wanes over time, these are all using relatively short-duration studies; vaccination has only been widely available since December 2020 so measures of efficacy are all truncated at 6-9 months. Whilst the available data can allow us to assess trends in the decline of vaccine efficacy, the long-term (asymptotic) levels are unknown. We therefore consider three levels of asymptotic protection, corresponding to 0%, 30% and 50% vaccine efficacy against infection (Figure 1); these are amalgamated in the main paper but are separated in the Supplementary Material. Protection against hospital admission and death is also assumed to wane, but to a higher asymptotic level such that severe outcomes are reduced by 70% conditional on infection compared with someone who has not been vaccinated.

Finally, although there is good data on antibody response following a booster dose [30], it is unclear precisely how this and other correlates of immunity will translate into long-term protection against infection and more severe outcomes [31]. We therefore model two extreme assumptions: either the third dose generates high and relatively long-lasting protection that wanes over slower timescales comparable to natural infection; or the third dose boosts protection to levels slightly higher than after the second dose which then wane over time in a similar manner to the waning after second doses (Figure 1 and Supplementary Material). Such boosting applies to all individuals whose protection (either following infection or vaccination) has waned, although this is dominated by individuals who have had two doses of vaccine and have not been infected.

Projections from October 2021 to October 2023 are shown in Figure 2, and focus on the number of daily hospital admissions due to COVID-19 (similar projections for deaths are given in the Supplementary Material). The historic model results (black line) are in excellent agreement with the data for England (red dots) showing the accuracy of the fitting mechanisms. Projecting these dynamics into the future shows that without boosters (dark blue line in all graphs) we would expect a large outbreak and associated hospitalisations in early 2022 surpassing the January 2021 second wave (which peaked at 4134 admissions), although the 95% prediction intervals are wide (95% prediction intervals capture the range of three assumptions about asymptotic protection and the impact of parameter uncertainty, both inferred and assumed, on the projected dynamics). We note that the three assumptions about asymptotic levels of protection require their own inferred parameters, constraining the model to agree with recent data despite the different levels of population immunity generated by the assumptions.

**Figure 2.**
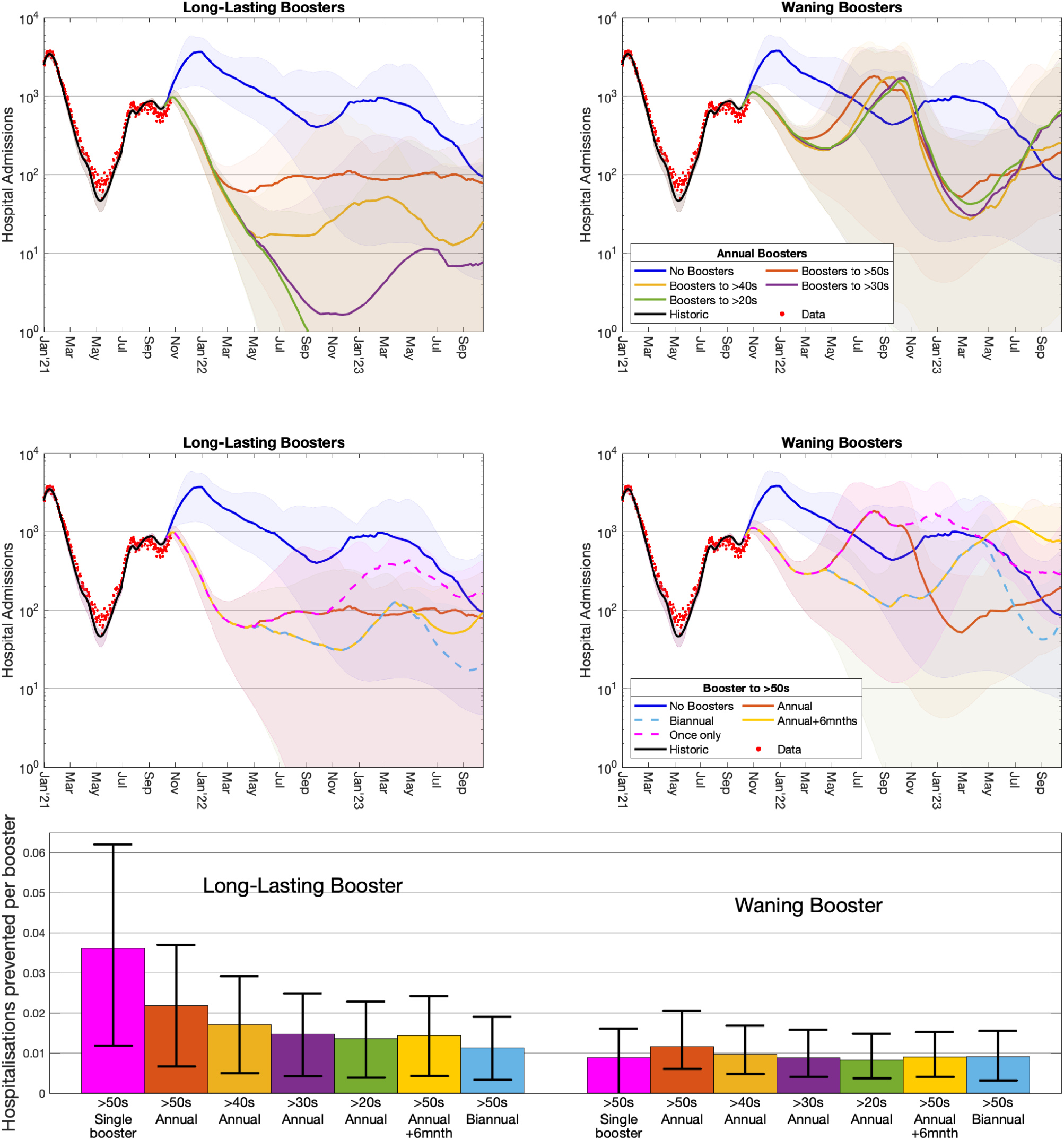
Comparison of the impact of boosters on hospital admissions per day to the counterfactual of no boosters. The top four graphs show projected hospital admissions per day in England over time (on a logarithmic scale) with data (red dots); the left-hand graphs are for the longer-lasting booster assumption while the right-hand graphs are for waning boosters. The top graphs show annual boosters (starting in September each year) given to different age-groups; the middle graphs show different temporal patterns of boosters given to the over fifties. The lower graph shows the number of hospital admissions prevented per booster dose (means and 95% prediction intervals) for the seven booster strategies considered relative to not offering boosters.

The introduction of boosters, at 1.3 million per week and at least 6 months after the second dose, suppresses much of the early 2022 wave. However, the longer-term dynamics are critically dependent on the protective characteristics following the booster. We assume a high uptake of the booster vaccine, with 95% of those over 70 years old and 90% of those under 70 who have received the first two doses of vaccine taking the third. When boosters offer high and relatively long-lasting efficacy (left column of Figure 2) extending the annual booster campaign to younger age-groups pushes future waves to be later and far smaller (comparing the red, yellow, purple and green lines). In contrast, when boosters wane at a similar speed to second-dose efficacy (right column of Figure 2), then the merits of extending an annual (autumn) booster campaign to younger age-groups is marginal, pushing subsequent waves to later in 2022 but doing little to mitigate overall or peak hospital admissions and only marginally reducing deaths. Under these pessimistic assumptions about the action of boosters, the use of biannual boosting every six months to the over 50s can truncate the larger waves, keeping the mean hospital admissions below a thousand per day for the entire time-frame simulated. Other patterns of boosters (such as adding a single six-month booster to the annual cycle, or only delivering a single round of boosters) lead to subsequent large waves in later years; however, under the longer-lasting assumption, a single round of boosters may be sufficient to suppress large numbers of hospital admissions for multiple years.

One way to quantify the projected impact of boosters is to calculate the average number of hospital admissions prevented by one booster (Figure 2 lower graph). For longer-lasting boosters, the maximal benefit per dose for annual boosters occurs when they are given to those over 50 years old; while offering boosters to younger age groups reduces the projected epidemic impact, the benefit per dose is less. The overall maximal benefit per dose occurs for a single booster given to the over 50s in Autumn 2021. This pattern is also seen in lives saved per booster (Supplementary Material). For projections in which boosters wane more rapidly, annual vaccination of over 50s generates the greatest number of hospital admissions prevented per dose; although a single booster in Autumn 2021 results in the greatest number of lives saved per booster dose.

Taken together these results highlight the need for high uptake of booster doses with a rapid delivery schedule (see Supplementary Material) as well as the vital importance of continual epidemiological monitoring to determine the level of population immunity across different age-groups [32]. The difference between rapidly waning immunity following boosters and much slower waning is stark, and determines the optimal path to endemicity, in particular whether regular boosters will be needed. Although we have considered the number of hospital admissions averted per booster dose as a single quantitative measure, there are multiple practical tradeoffs in terms of minimising the pressure on health services and hence reducing peak hospital admissions. The chosen booster program of targeting the over 50s, healthcare workers and clinically vulnerable appears optimal in the short term, but may need to be supplemented by additional boosters in these at-risk groups if efficacy is observed to decline.

## Data Availability

The ethics of the use of these data for these purposes was agreed by the UK Health Security Agency with the Governments SPI-M(O) / SAGE committees.

## Author contributions

**Matt J. Keeling:** Conceptualisation, Data curation, Formal analysis, Funding acquisition, Methodology, Software, Visualisation, Writing - Original Draft, Writing - Review & Editing.

**Amy Thomas:** Methodology, Visualisation, Writing - Original Draft, Writing - Review & Editing.

**Edward M. Hill:** Methodology, Visualisation, Writing - Review & Editing.

**Robin N. Thompson:** Methodology, Visualisation, Writing - Review & Editing.

**Louise Dyson:** Funding acquisition, Methodology, Visualisation, Writing - Review & Editing.

**Michael J. Tildesley:** Funding acquisition, Methodology, Visualisation, Writing - Review & Editing.

**Sam Moore:** Methodology, Visualisation, Writing - Review & Editing.

## Data availability

The ethics of the use of these data for these purposes was agreed by the UK Health Security Agency with the Government’s SPI-M(O) / SAGE committees.

## Footnotes

1 We note that achieving an estimate of percentage uptake is confounded by needing a robust estimate of the population size in each age-group.

## References

1. Woolf SH, Chapman DA, Lee JH. COVID-19 as the Leading Cause of Death in the United States. JAMA. 2021;325: 123–124. doi:10.1001/jama.2020.24865

2. Woolf SH, Chapman DA, Sabo RT, Weinberger DM, Hill L. Excess Deaths From COVID-19 and Other Causes, March-April 2020. JAMA. 2020;324: 510–513. doi:10.1001/jama.2020.11787

3. Simonsen L, Viboud C. A comprehensive look at the COVID-19 pandemic death toll. Elife. 2021;10: e71974. doi:10.7554/eLife.71974

4. Karlinsky A, Kobak D. Tracking excess mortality across countries during the COVID-19 pandemic with the World Mortality Dataset. Elife. 2021;10: e69336. doi:10.7554/eLife.69336

5. Office for National Statistics. Coronavirus and the impact on output in the UK economy: August 2021. 2021. Available: https://www.ons.gov.uk/economy/grossdomesticproductgdp/articles/coronavirusandtheimpactonoutputintheukeconomy/august2021

6. Giuntella O, Hyde K, Saccardo S, Sadoff S. Lifestyle and mental health disruptions during COVID-19. Proc Natl Acad Sci. 2021;118: e2016632118. doi:10.1073/pnas.2016632118

7. Levin EG, Lustig Y, Cohen C, Fluss R, Indenbaum V, Amit S, et al. Waning Immune Humoral Response to BNT162b2 Covid-19 Vaccine over 6 Months. N Engl J Med. 2021. doi:10.1056/NEJMoa2114583

8. Khoury DS, Cromer D, Reynaldi A, Schlub TE, Wheatley AK, Juno JA, et al. Neutralizing antibody levels are highly predictive of immune protection from symptomatic SARS-CoV-2 infection. Nat Med. 2021;27: 1205–1211. doi:10.1038/s41591-021-01377-8

9. Volz E, Mishra S, Chand M, Barrett JC, Johnson R, Geidelberg L, et al. Assessing transmissibility of SARS-CoV-2 lineage B.1.1.7 in England. Nature. 2021;593: 266–269. doi:10.1038/s41586-021-03470-x

10. Davies NG, Abbott S, Barnard RC, Jarvis CI, Kucharski AJ, Munday JD, et al. Estimated transmissibility and impact of SARS-CoV-2 lineage B.1.1.7 in England. Science. 2021;372: eabg3055. doi:10.1126/science.abg3055

11. Kraemer MUG, Hill V, Ruis C, Dellicour S, Bajaj S, McCrone JT, et al. Spatiotemporal invasion dynamics of SARS-CoV-2 lineage B.1.1.7 emergence. Science. 2021; eabj0113. doi:10.1126/science.abj0113

12. Sonabend R, Whittles LK, Imai N, Perez-Guzman PN, Knock ES, Rawson T, et al. Non-pharmaceutical interventions, vaccination, and the SARS-CoV-2 delta variant in England: a mathematical modelling study. Lancet. 2021. doi:10.1016/S0140-6736(21)02276-5

13. Allen H, Vusirikala A, Flannagan J, Twohig KA, Zaidi A, Chudasama D, et al. Household transmission of COVID-19 cases associated with SARS-CoV-2 delta variant (B.1.617.2): national case-control study. Lancet Reg Heal – Eur. 2021. doi:10.1016/j.lanepe.2021.100252

14. Baraniuk C. Covid-19: How the UK vaccine rollout delivered success, so far. BMJ. 2021;372: n421. doi:10.1136/bmj.n421

15. Moore S, Hill EM, Dyson L, Tildesley MJ, Keeling MJ. Modelling optimal vaccination strategy for SARS-CoV-2 in the UK. PLOS Comput Biol. 2021;17: e1008849. Available: https://doi.org/10.1371/journal.pcbi.1008849

16. UK Health Security Agency. COVID-19 vaccine weekly surveillance reports. 2021. Available: https://www.gov.uk/government/publications/covid-19-vaccine-weekly-surveillance-reports

17. Stephens DS, McElrath MJ. COVID-19 and the Path to Immunity. JAMA. 2020;324: 1279–1281. doi:10.1001/jama.2020.16656

18. Suthar MS, Zimmerman MG, Kauffman RC, Mantus G, Linderman SL, Hudson WH, et al. Rapid Generation of Neutralizing Antibody Responses in COVID-19 Patients. Cell Reports Med. 2020;1: 100040. doi:https://doi.org/10.1016/j.xcrm.2020.100040

19. Lumley SF, Wei J, O’Donnell D, Stoesser NE, Matthews PC, Howarth A, et al. The Duration, Dynamics, and Determinants of Severe Acute Respiratory Syndrome Coronavirus 2 (SARS-CoV-2) Antibody Responses in Individual Healthcare Workers. Clin Infect Dis. 2021;73: e699–e709. doi:10.1093/cid/ciab004

20. Dan JM, Mateus J, Kato Y, Hastie KM, Yu ED, Faliti CE, et al. Immunological memory to SARS-CoV-2 assessed for up to 8 months after infection. Science. 2021;371: eabf4063. doi:10.1126/science.abf4063

21. Xia W, Li M, Wang Y, Kazis LE, Berlo K, Melikechi N, et al. Longitudinal analysis of antibody decay in convalescent COVID-19 patients. Sci Rep. 2021;11: 16796. doi:10.1038/s41598-021-96171-4

22. Naaber P, Tserel L, Kangro K, Sepp E, Jürjenson V, Adamson A, et al. Dynamics of antibody response to BNT162b2 vaccine after six months: a longitudinal prospective study. Lancet Reg Heal - Eur. 2021; 100208. doi:10.1016/j.lanepe.2021.100208

23. Office for National Statistics. Coronavirus (COVID-19) Infection Survey technical article: analysis of reinfections of COVID-19: June 2021. 2021. Available: https://www.ons.gov.uk/peoplepopulationandcommunity/healthandsocialcare/conditionsanddiseases/articles/coronaviruscovid19infectionsurveytechnicalarticleanalysisofreinfectionsofcovid19/june2021

24. Hansen CH, Michlmayr D, Gubbels SM, Mølbak K, Ethelberg S. Assessment of protection against reinfection with SARS-CoV-2 among 4 million PCR-tested individuals in Denmark in 2020: a population-level observational study. Lancet. 2021;397: 1204–1212. doi:10.1016/S0140-6736(21)00575-4

25. Andrews N, Tessier E, Stowe J, Gower C, Kirsebom F, Simmons R, et al. Vaccine effectiveness and duration of protection of Comirnaty, Vaxzevria and Spikevax against mild and severe COVID-19 in the UK. medRxiv. 2021; 2021.09.15.21263583. doi:10.1101/2021.09.15.21263583

26. Pouwels KB, Pritchard E, Matthews PC, Stoesser N, Eyre DW, Vihta K-D, et al. Effect of Delta variant on viral burden and vaccine effectiveness against new SARS-CoV-2 infections in the UK. Nat Med. 2021. doi:10.1038/s41591-021-01548-7

27. Keeling MJ, Hill EM, Gorsich EE, Penman B, Guyver-Fletcher G, Holmes A, et al. Predictions of COVID-19 dynamics in the UK: Short-term forecasting and analysis of potential exit strategies. Flegg JA, editor. PLOS Comput Biol. 2021;17: e1008619. doi:10.1371/journal.pcbi.1008619

28. Dyson L, Hill EM, Moore S, Curran-Sebastian J, Tildesley MJ, Lythgoe KA, et al. Possible future waves of SARS-CoV-2 infection generated by variants of concern with a range of characteristics. Nat Commun. 2021;12: 5730. doi:10.1038/s41467-021-25915-7

29. Moore S, Hill EM, Tildesley MJ, Dyson L, Keeling MJ. Vaccination and non-pharmaceutical interventions for COVID-19: a mathematical modelling study. Lancet Infect Dis. 2021;21: 793–802. doi:10.1016/S1473-3099(21)00143-2

30. Flaxman A, Marchevsky NG, Jenkin D, Aboagye J, Aley PK, Angus B, et al. Reactogenicity and immunogenicity after a late second dose or a third dose of ChAdOx1 nCoV-19 in the UK: a substudy of two randomised controlled trials (COV001 and COV002). Lancet. 2021;398: 981–990. doi:10.1016/S0140-6736(21)01699-8

31. Bar-On YM, Goldberg Y, Mandel M, Bodenheimer O, Freedman L, Kalkstein N, et al. Protection of BNT162b2 Vaccine Booster against Covid-19 in Israel. N.Engl.J.Med. 2021; 385:1393–1400 doi:10.1056/NEJMoa2114255

32. Office for National Statistics. Coronavirus (COVID-19) Infection Survey, UK Statistical bulletins. 2021. Available: https://www.ons.gov.uk/peoplepopulationandcommunity/healthandsocialcare/conditionsanddiseases/bulletins/coronaviruscovid19infectionsurveypilot/previousReleases

