## Supplementary Information for "Waning, Boosting and a Path to Endemicity for SARS-CoV-2"

#### Model Structure and the Action of Boosters

The modelling of booster vaccination outlined in the main paper is based upon the mathematical model of SARS-CoV-2 infection and COVID-19 disease that has been developed in the University of Warwick since early 2020. The model has been used in a number of settings [1,2] and has been described in detail in multiple publications [3,4]; here we provide a synopsis of the model structure and focus on the new elements that are key to modelling waning efficacy and immunity as well as the action of booster vaccines.

We use a compartmental, deterministic age-structured model, developed to simulate the spread of SARS-CoV-2 within seven regions in England (corresponding to NHS regions: East of England, London, Midlands, North East & Yorkshire, North West, South East and South West), with parameters inferred to generate a fit to deaths, hospital admissions, hospital occupancy, ICU occupancy, proportion of tests that are positive and serological testing [5]. The model population is stratified into 5-year age classes (0-4yrs, 5-9yrs, …, 95-99yrs, 100+yrs), with the force of infection determined by the use of an age-dependent (who acquires infection from whom) social contact matrix for the UK [35]. Additionally, we allow susceptibility and the probabilities of becoming symptomatic, being hospitalised and the risk of dying to be age dependent; these are all matched to UK outbreak data. Finally, we account for the role of household isolation, by separating primary and secondary infections within a household (more details may be found in [3,5]). This allows us to capture household isolation by preventing secondary infections from playing a further role in onward transmission. Model parameters were inferred on a regional basis matching to epidemiological observations in the seven NHS regions of England. More details of the model structure are provided at the end of the Supplementary material, but here we focus on the novel aspects of third dose vaccine boosting (Figure S1).

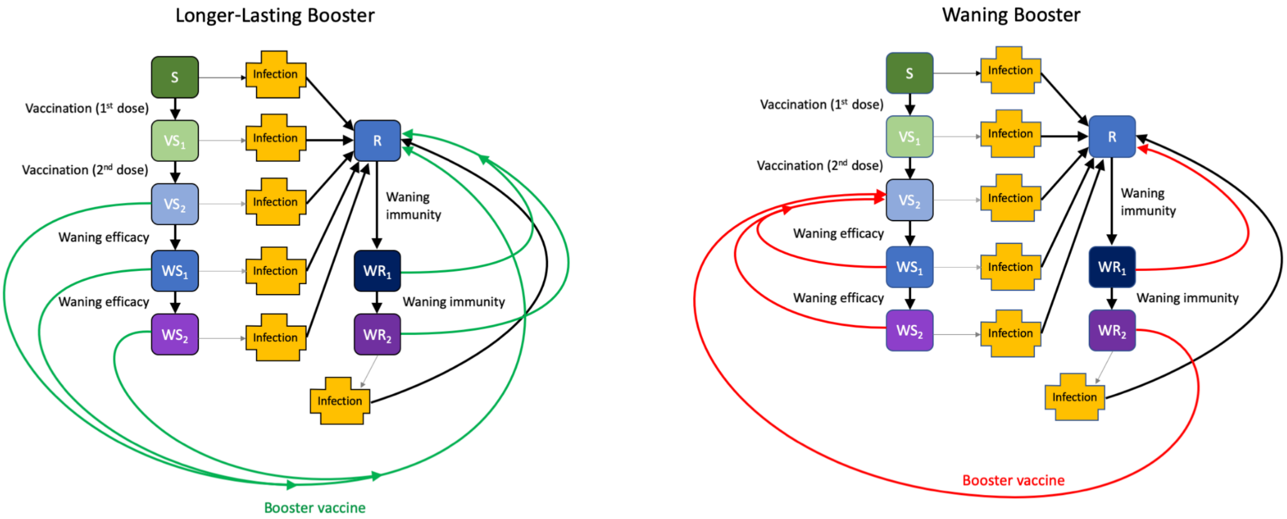


**Figure S1**. **Two alternative models of vaccine booster doses**, that give rise to longer-lasting boosters (left) and repeated waning boosters (right). Here the complex infection dynamics have been encapsulated within a single box, and we focus on the transitions of non-infected individuals. All individuals begin the simulations as susceptible and move through the vaccinated classes ($VS_1$ and $VS_2$) on receipt of first and second doses of the vaccine although this eventually wanes as part of a two-step process ($WS_1$ and $WS_2$); those that recover from infection (class R) also experience waning immunity (classes $WR_1$ and $WR_2$), although over slower time-scales. For longer-lasting immunity (left-hand diagram) the booster vaccine provides similar protection and over a similar time scale as recovery from infection. In contrast, for waning boosters the third dose takes waned individuals back to the 'double vaccinated' state ($VS_2$); the only exception is those in class $WR_1$ (very early stages of waning after recovery) who return to the R class.

The pathway for non-infected individuals is governed by vaccination and waning. Susceptible individuals transition into different classes on receiving their first and second dose of vaccine; individuals in these classes have a lower risk of infection and, if infected, a lower risk of severe outcomes (hospital admission or death) - see Table 1.

Individuals wane from these vaccinated states in a two-step process to capture the observed decay in vaccine efficacy. Efficacy in state $WS_2$ is lower than in $VS_2$ and the asymptotic waning efficacy against infection is assumed to take one of three values: VE → 50%; VE → 30%; or VE → 0%, where VE → 50% corresponds to limited decay in efficacy beyond what has already been observed at six months after the second dose of vaccine whilst VE → 0% corresponds to an eventual complete loss of protection against infection. In this waning state, there is still a 70% reduction in severe disease following infection, compared with naive susceptible individuals.

**Table 1**. Efficacy for two doses of Pfizer (BNT162b2) and AstraZeneca (ChAdOx1) vaccine and for the three waning efficacy assumptions used throughout the paper. The term in brackets in the top row shows the compartments in Figure S1 where the VE assumptions apply. For the waning assumptions we also give the mean time to reach this reduced efficacy from deployment of the second dose.

| VE against | Following infection (R, $WR_1$) | Pfizer 2 doses ($VS_2$, $WS_1$) | AZ 2 doses ($VS_2$, $WS_1$) | Assumption VE → 50% ($WS_2$, $WR_2$) | Assumption VE → 30% ($WS_2$, $WR_2$) | Assumption VE → 0% ($WS_2$, $WR_2$) |
|---|---|---|---|---|---|---|
| Infection | 100% | 85% | 70% | 50% | 30% | 0% |
| Symptoms | 100% | 90% | 75% | 55% | 35% | 5% |
| Hospital Adm. | 100% | 95% | 95% | 85% | 79% | 70% |
| Death | 100% | 98% | 98% | 85% | 79% | 70% |
| Waning time (mean days to asymptote) | R→$WR_2$ 1860 | - | - | $VS_2$→$WS_2$ 180 | $VS_2$→$WS_2$ 310 | $VS_2$→$WS_2$ 460 |

The average times in states $VS_2$ and $WS_1$ (where efficacy is identical to being in state $VS_2$) are dependent on the level of asymptotic efficacy and are parameterised to match the efficacy values estimated at different times from second dose (Table 1).

We make the simplifying assumption that the immunity gained from infection dominates any immunity from vaccination. Thus, individuals that are recovered from infection and receive the vaccine remain in the recovered class (R in Figure S1); similarly, vaccinated individuals who get infected and recover are placed in the same recovered class. The immunity derived from infection also wanes, such that recovered individuals follow a similar path of waning as vaccinated individuals (shown as R, $WR_1$ and $WR_2$ in Figure S1). Waning of infection-derived immunity is assumed to wane more slowly than vaccine efficacy (with mean time to progress from R to $WR_2$ approximately 5 years) in keeping with the observation that re-infection events remain relatively uncommon. In the absence of other data we assume that the asymptotic level of protection is the same as for waning after vaccination, following the three assumptions in Table 1. Over the time-scales simulated here, our results are not highly sensitive to this speed of waning nor the asymptotic level associated with recovered individuals, as these parameters are compensated for within the inference framework to enable good fit to the data.

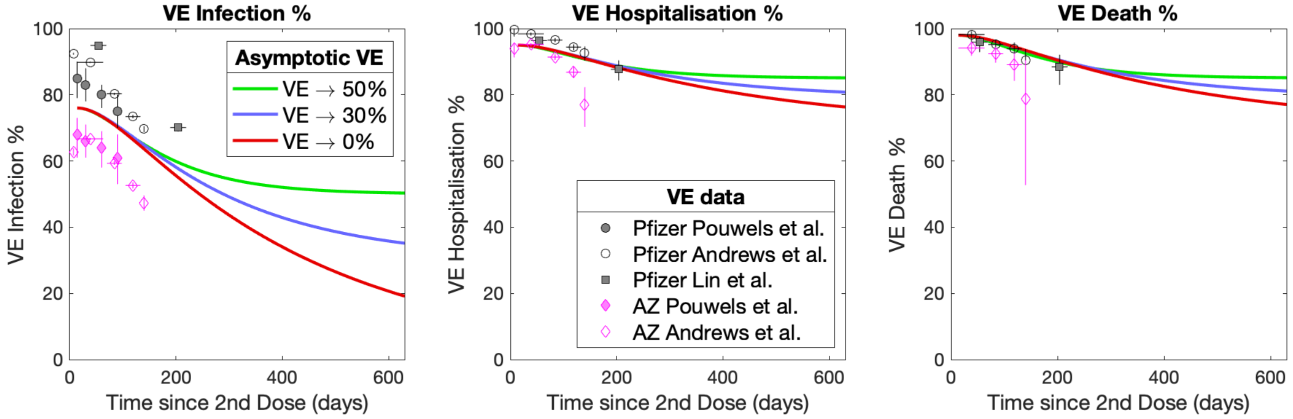


**Figure S2**. **Comparison between model assumptions (curves) and data [6-8]** for the three different measures of vaccine efficacy (against infection, hospital admissions and deaths). The curves represent the average efficacy from those in compartments $VS_2$, $WS_1$, $WS_2$ where 50% of those in $VS_2$ (which sets the early efficacy) are assumed to have the AstraZeneca (ChAdOx1) vaccine, while the remainder had an mRNA vaccine considered to have the same characteristics of Pfizer (BNT162b2). The data point show 95% confidence intervals and range of times since second dose from three studies [6-8], noting that [8] is from the USA while [6,7] are from the UK.

Booster doses (shown as red or green arrows in Figure S1) act to overcome waning and increase individual-level protection. Throughout the paper, we consider two assumptions for the action of booster doses, labelled longer-lasting boosters and waning boosters. Longer-lasting boosters (left-hand diagram) assume that boosters have the same effect as recovery from infection and generate strong and relatively long-lasting immunity. In contrast, for the rapidly waning assumption, boosters effectively act in a similar manner to second doses of vaccine, generating high levels of immunity that wane relatively quickly (Table 2).

**Table 2**. Efficacy following the third booster dose, either generating longer-lasting immunity or waning boosters. When the booster wanes rapidly, the three time-scales are associated with the three assumptions about efficacy in the waned state (VE → 50%, VE → 30%, and VE → 0%).

| VE against | Longer-lasting boosters | Waning boosters |
|---|---|---|
| Infection | 100% | 92% |
| Symptoms | 100% | 95% |
| Hospital Adm. | 100% | 97% |
| Death | 100% | 99% |
| Waning time (mean days to asymptote) | 1860 | 180, 310, 460 |

In total we therefore consider six different combinations of assumptions that determine long-term dynamics: three that investigate the impact of varying the asymptotic level of vaccine efficacy and two that explore the action of boosters.

**Implication of Boosters for COVID-19 Deaths**

Here we show the projected number of daily deaths in England until October 2023. As such this is derived from the same simulations as Figure 2 in the main paper and reflects the eight booster scenarios considered. In general, the results echo the findings for hospitalisations, with longer-lasting boosters able to drive deaths to a relatively low level but when boosters wane rapid additional booster programmes may be needed.

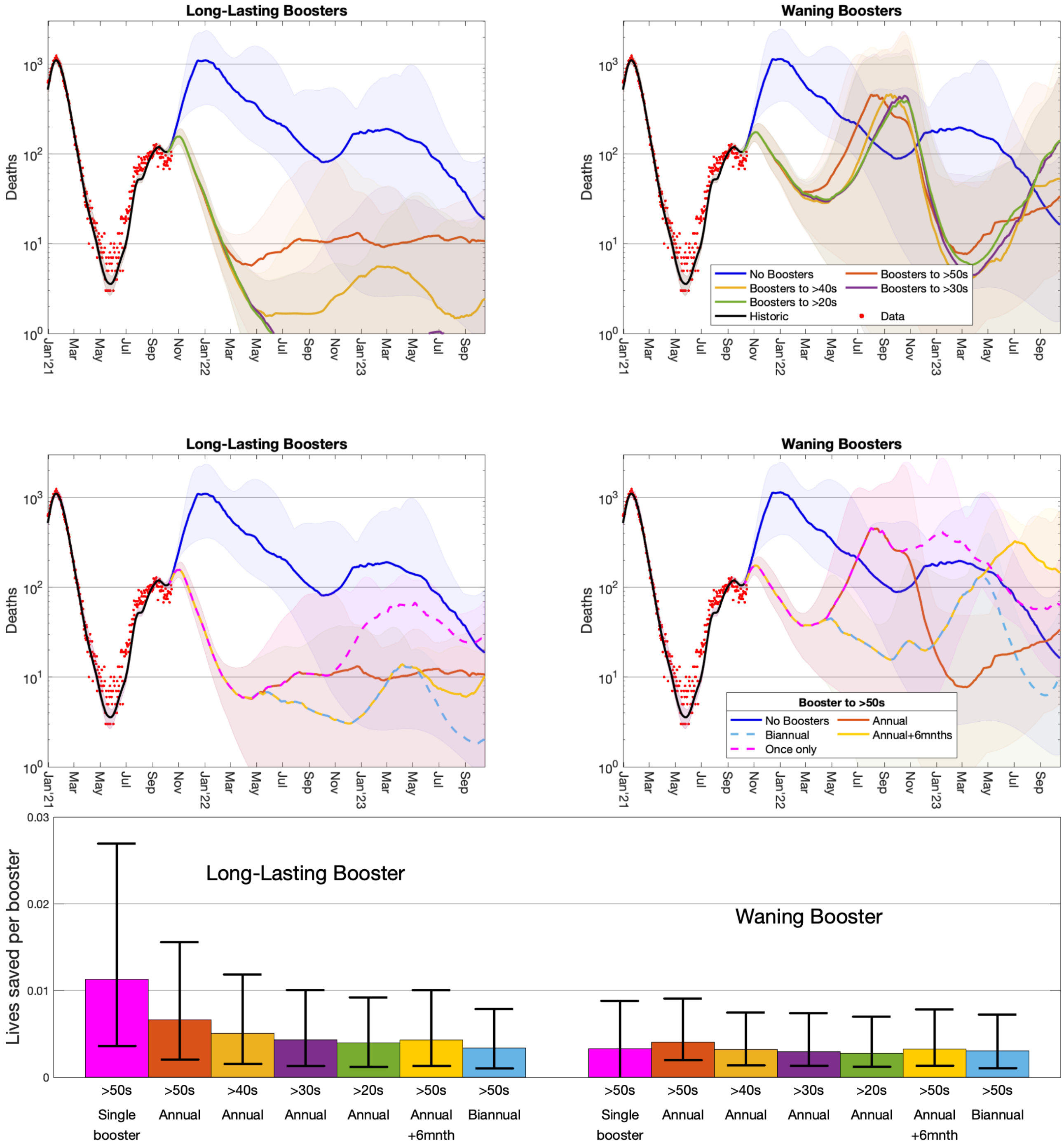


**Figure S3 Comparison of the impact on deaths of boosters to the counterfactual of no boosters.** The top four graphs show projected daily deaths in England over time (on a logarithmic scale) with data (red dots); the left-hand graphs are for the longer-lasting booster assumption while the right-hand graphs are for waning boosters. The lower graph shows the number of lives saved per booster dose (means and 95% prediction intervals) for the seven booster strategies considered relative to not offering boosters.

### Implication of Boosters for COVID-19 Hospital Admissions (linear scale)

Throughout we have shown all model projections on a logarithmic scale, as this more accurately allows visualisation of all results which can differ by orders of magnitude. However, the pressure on public health services is clearly related to the number of hospital admissions which can be assessed more easily on a linear scale. This is presented in Figure S4, which is the same as Figure 2 in the main text but shown on a linear scale.

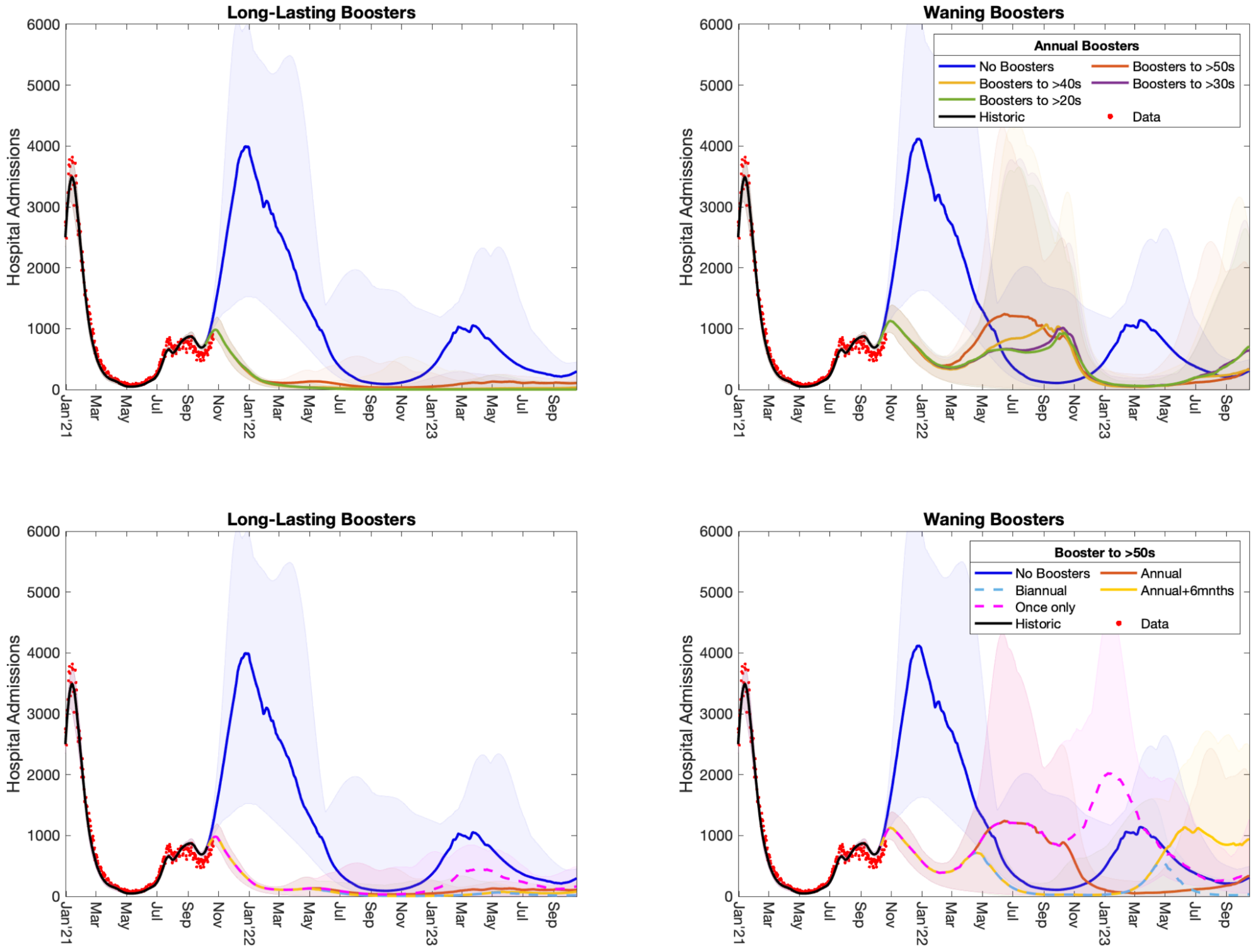


**Figure S4. Comparison of the impact of boosters on hospital admissions per day to the counterfactual of no boosters.** The four graphs show projected hospital admissions per day in England over time (on a linear scale) with data (red dots); the left-hand graphs are for the longer-lasting booster assumption while the right-hand graphs are for waning boosters. The top graphs show annual boosters (starting in September each year) given to different age-groups, the lower graphs show different temporal patterns of boosters given to the over fifties.

**Separating the heterogeneity in assumptions about waning efficacy.**

In the main paper, to reduce the combinatorial dimension of the uncertainty, the results for the three waning assumptions (where VE→0%, VE→30%, VE→50%, see Figure 1 of the main text) were combined. Here we separate this additional uncertainty, with Figure S5 showing the number of hospital admissions in England under the assumption of long-lasting boosters, and Figure S6 showing the results for the more rapid waning of booster assumption.

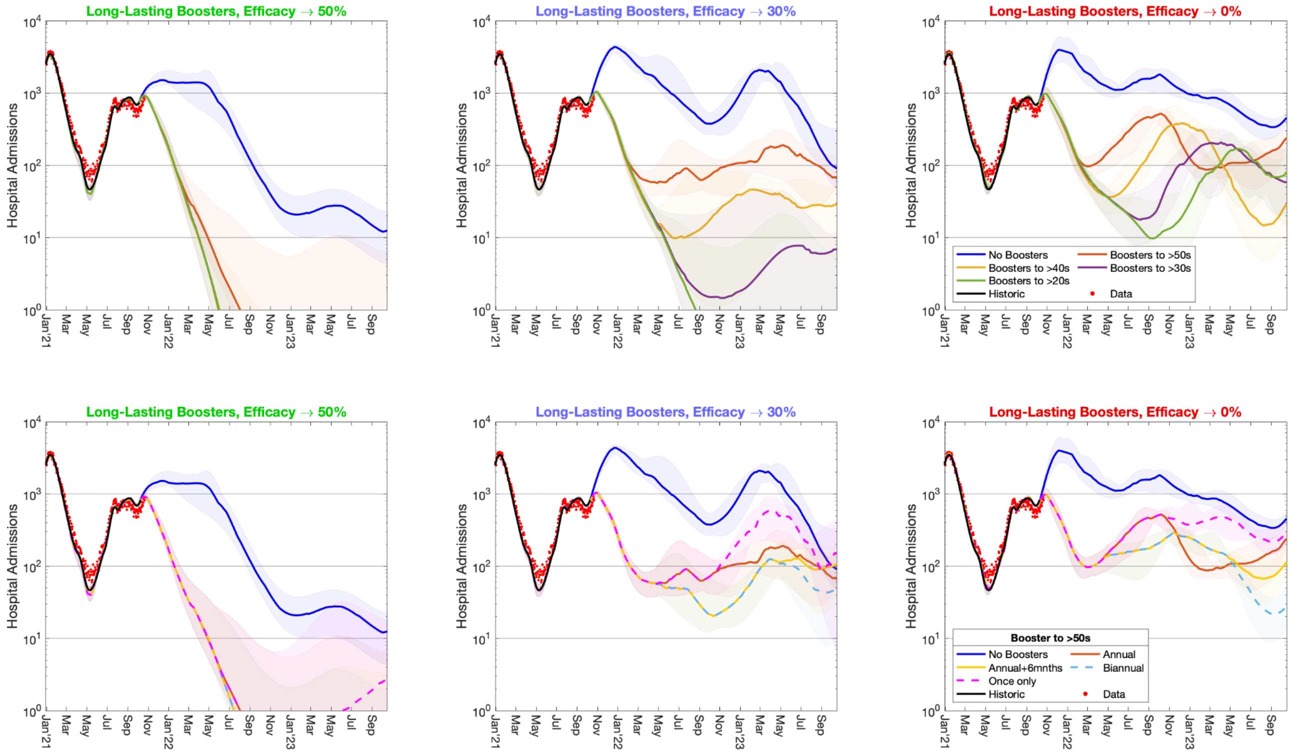


**Figure S5 Comparison of the impact of boosters to the counterfactual of no boosters for the longer-lasting booster assumption**. The four graphs show projected daily hospital admissions in England over time (on a logarithmic scale) with data (red dots); the left-hand graphs are for the optimistic assumption where vaccine efficacy against infection only wanes to 50%, the central graphs are when this efficacy wanes to 30%, and the right-hand graphs are for the pessimistic case where vaccine efficacy against infection wanes to 0%. As shown in Figure 1 of the main text, the waning time associated with each of these assumptions is tuned to match the currently available data.

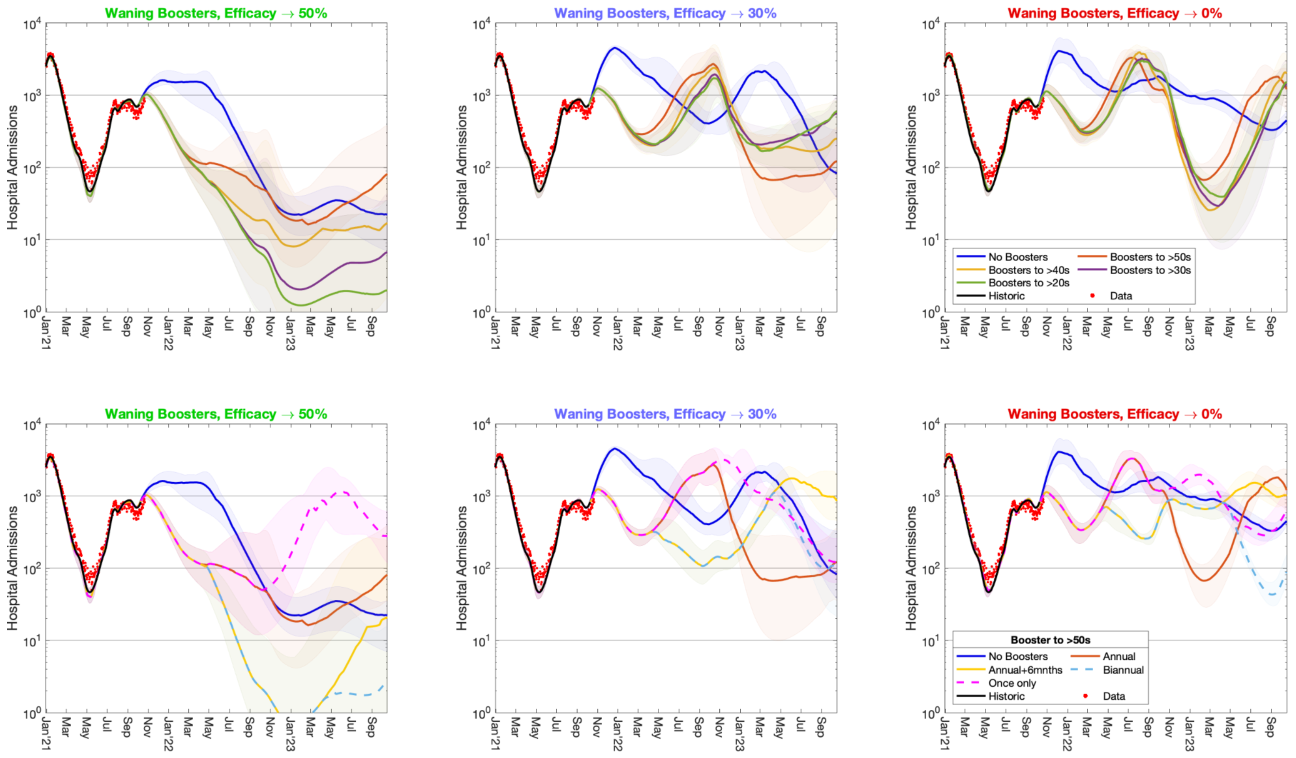


**Figure S6 Comparison of the impact of boosters to the counterfactual of no boosters for the assumption where the efficacy from boosters wane** in a similar manner to the efficacy from two doses of vaccine. The four graphs show projected daily hospital admissions in England over time (on a logarithmic scale) with data (red dots); the left-hand graphs are for the optimistic assumption where vaccine efficacy against infection only wanes to 50%, the central graphs are when this efficacy wanes to 30%, and the right-hand graphs are for the pessimistic case where vaccine efficacy against infection wanes to 0%. As shown in Figure 1 of the main text, the waning time associated with each of these assumptions is tuned to match the currently available data

**Incremental Benefits per Booster**

In the main text we presented estimates of the number of hospital admissions prevented per booster dose. Here we adapt the concept of incremental cost-effectiveness analysis to consider the additional benefit from expanding each booster vaccination programme (Figure S7). We start with the current (minimal) strategy of a single booster dose being offered to all adults over 50 (pink), which is compared with the counterfactual of no booster vaccine. Subsequent extensions of this strategy are then compared to the previous largest strategy (such as the move to annual vaccination of over 50s (orange) is compared to the single dose to over 50s (pink), while the annual booster vaccination of over 40s (yellow) is compared to the annual booster vaccination of over 50s). As such each bar quantifies the benefit (in terms of hospital admission or deaths prevented per booster dose) of each increment in the booster programme.

Strategies written in bold have a statistically significant advantage over preceding strategies (the lower bound of the 95% prediction interval is greater than zero). For the longer-lasting booster, the single booster campaign to the over 50s is hugely beneficial, with some very weak support for annual boosters to everyone over 40. For the assumption where the efficacy from boosters wanes more rapidly, annual vaccination for the over 50s gets the strongest support, with some weak support for either annual boosting of the over 40s or adding an additional booster for the over 50s after another six months.

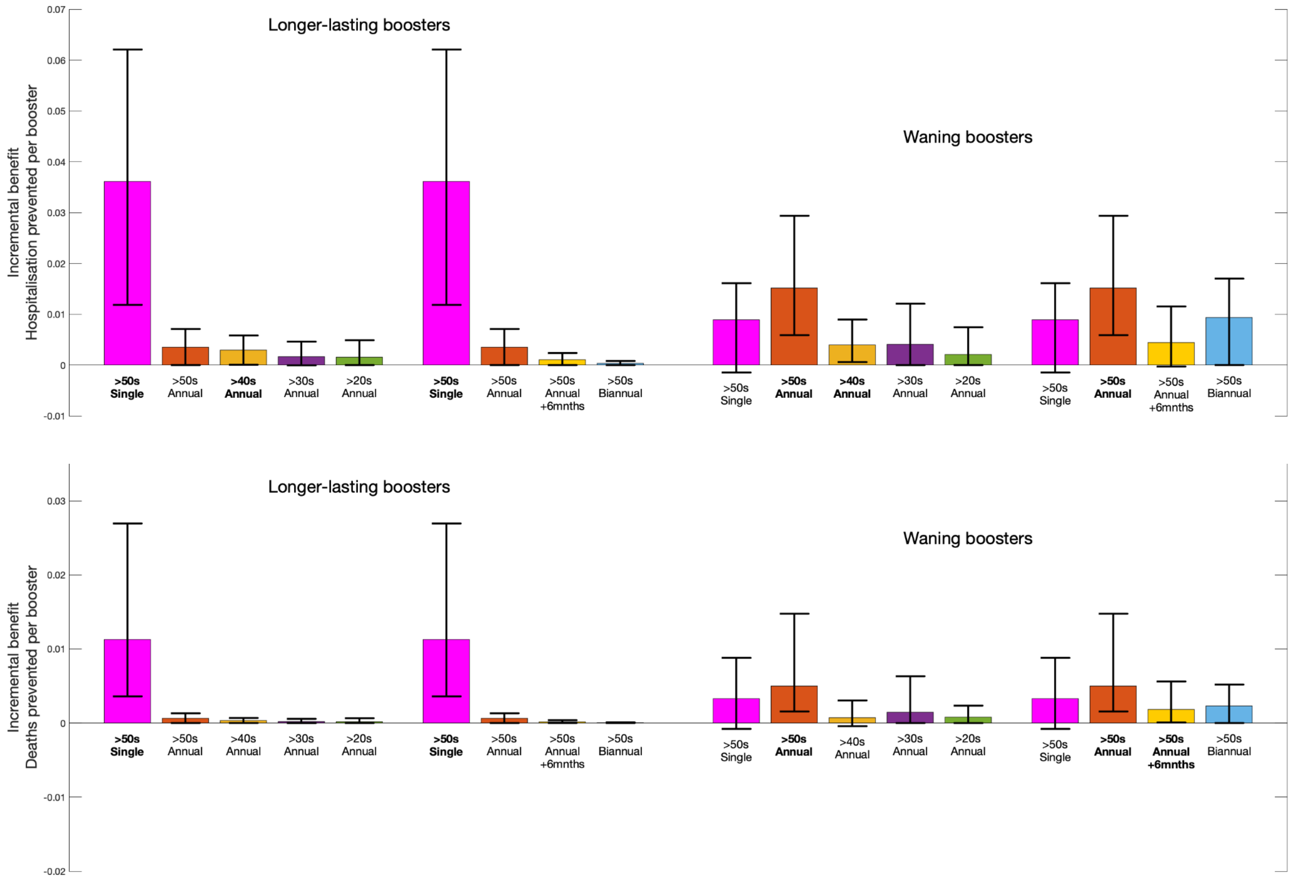


**Figure S7. Incremental Benefit Analysis of booster vaccinations against hospital admissions (top) and deaths (bottom).** The left-hand set of bars is for longer-lasting boosters, the right-hand set is for more rapidly waning boosters. Each bar represents the additional benefit per additional vaccine dose of extending the programme to include a large amount of vaccination (compared to the bar to the left) - the single dose booster given to the over 50s is compared to not giving any boosters. Strategies written in bold have a statistically significant benefit over less intensive strategies.

It is worth stressing two important points about the analysis in Figure S7. Firstly, it only considers the total number of hospital admissions (or deaths) over the period October 2021 to October 2023; it does not account for peaks in admission which may place the health system under more stress than a prolonged wave of similar numbers. Secondly, there are no costs in this analysis, only benefits per dose; the move toward endemicity may necessitate a more traditional cost-benefit analysis where the costs of the vaccine are compared to the costs of treatment and the associated quality of life (QALY) impacts [9-13]

**Effects of lower booster uptake.**

Throughout this manuscript we have assumed that uptake of boosters by anyone that has already received two doses of vaccine will be high: 95% for those over 70 years of age, and 90% for everyone else. In Figure S8, we consider relative changes to this uptake level for annual boosting of the over 50s; as such 100% relative uptake corresponds to 95% for those over 70 and 90% for those under 70, whereas 50% relative uptake corresponds to 47.5% in the older age group and 45% in the younger age group. For comparison we also show no boosters (which can be considered as 0% uptake).

For long-lasting booster, there is a very pronounced impact of booster uptake with a large advantage for high uptake. For the waning booster the case is less clear; higher levels of uptake generate an advantage, but waves of infection in mid-2022 partially compensate for some of the early gains.

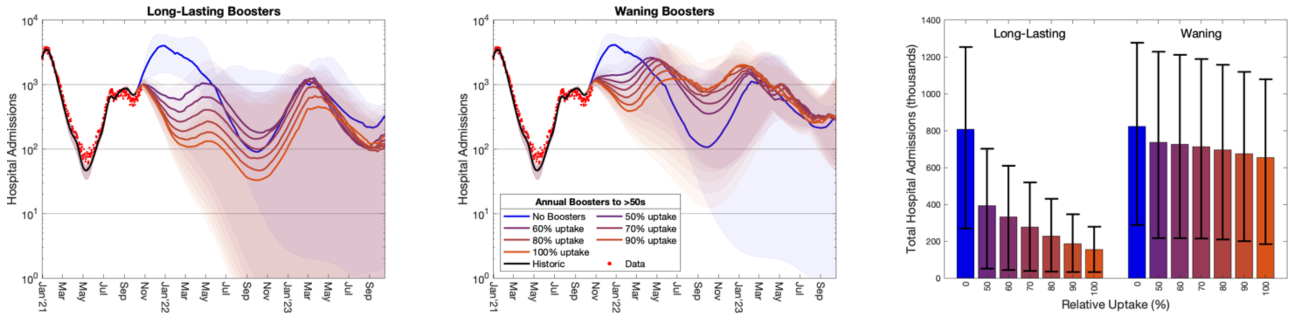


**Figure S8. Comparison of booster uptake on hospital admissions assuming annual boosters given to the over 50s.** The left and centre graphs show projected hospital admissions per day in England over time (on a logarithmic scale) with data (red dots); the left-hand graph is for the longer-lasting booster assumption while the centre graph is for waning boosters. The right-hand graph is the total number of projected hospital admissions from Oct 2021 to Oct 2023 for the two booster assumptions and seven levels of relative uptake (error bars show the 95% prediction intervals).

**Model Description**

Here we detail the underlying mathematical framework that defines the model. We break the model into multiple sections that combine to generate a picture of SARS-CoV-2 transmission in the UK. This model structure has already been detailed in previous publications [1-3] but we review the details here for completeness.

**Infection modelling**

As is common to most epidemiological modelling we stratify the population into multiple disjoint compartments and capture the flow of individuals between compartments in terms of ordinary differential equations. At the heart of the model is a modified SEIR equation, where individuals may be susceptible (S), exposed (E), infectious with symptoms (I), infectious and either asymptomatic or with very mild symptoms (A) or recovered (R). Both symptomatic and asymptomatic individuals are able to transmit infection, but asymptomatics do so at a reduced rate given by $\tau$. Hence the force of infection is proportional to I+$\tau$A. To some extent, the separation into symptomatic (I) and asymptomatic (A) within the model is somewhat artificial as there are a wide spectrum of symptom severities that can be experienced, with the classification of symptoms changing over time. Here our classification reflects early case detection, when only relatively severe symptoms were recognised. To obtain a better match to the infection time scales, we model the exposed class as a 3-stage process - this provides a better match to the time from infection to becoming infectious, such that in a stochastic formulation the distribution of the latent period would be an Erlang distribution.

$$
\begin{aligned}
\frac{dS}{dt} &= -\lambda S \qquad\qquad \text{where } \lambda \propto (I + \tau A) \\
\frac{dE_1}{dt} &= \lambda S - 3\alpha E_1 \\
\frac{dE_2}{dt} &= 3\alpha E_1 - 3\alpha E_2 \\
\frac{dE_3}{dt} &= 3\alpha E_2 - 3\alpha E_3 \\
\frac{dI}{dt} &= 3d\alpha E_3 - \gamma I \\
\frac{dA}{dt} &= 3(1-d)\alpha E_3 - \gamma A \\
\frac{dR}{dt} &= \gamma(I + A)
\end{aligned}
$$

where $\alpha^{-1}$, and $\gamma^{-1}$ are the mean latent and infectious periods, while *d* is the proportion of infections that develop symptoms.

**Age Structure & Transmission Structure**

The simple model structure is expanded to twenty-one 5-year age-groups (0-4, 5-9, .... ,95-99, 100+). Age has three major impacts on the epidemiological dynamics, with each element parameterised from the available data:

- Older individuals have a higher susceptibility to SARS-CoV-2 infection.
- Older individuals have a higher risk of developing symptoms, and therefore have a greater rate of transmission per contact.
- Older individuals have a higher risk of more severe consequences of infection including hospital admission and death.

The age-groups interact through four who-acquired-infection-from-whom transmission matrices, which capture the epidemiological relevant mixing in four settings: household ($\boldsymbol{\beta}^{H}$), school ($\boldsymbol{\beta}^{S}$),

workplace ($\boldsymbol{\beta}^{W}$) and other ($\boldsymbol{\beta}^{O}$). We took these matrices from Prem *et al.* [14] to allow easy translation to other geographic settings, although other sources such as POLYMOD [15] could be used.
One of the main modifiers of mixing and therefore transmission is the level of precautionary behaviour, $\phi$ (see Figure 1 of the main text). This changes the who-acquired-infection-from-whom transmission matrices in each transmission setting, such that when $\phi$=1 mixing in workplaces and other settings take their lowest value, whereas when $\phi$=0 the mixing returns to pre-pandemic levels. Mixing within the school setting follows the prescribed opening and closing of schools.

$$\begin{aligned}
\frac{dS_a}{dt} &= -\lambda_a S_a \qquad\qquad \text{where } \lambda_a = \sum_b \beta_{ab}(\phi)(I_b + \tau_b A_b) \\
\frac{dE_{a1}}{dt} &= \lambda S_a - 3\alpha E_{a1} \\
\frac{dE_{a2}}{dt} &= 3\alpha E_{a1} - 3\alpha E_{a2} \\
\frac{dE_{a3}}{dt} &= 3\alpha E_{a2} - 3\alpha E_{a3} \\
\frac{dI_a}{dt} &= 3 d_a \alpha E_{a3} - \gamma I_a \\
\frac{dA_a}{dt} &= 3(1 - d_a)\alpha E_{a3} - \gamma A_a \\
\frac{dR_a}{dt} &= \gamma(I_a + A_a)
\end{aligned}$$

For simplicity of notation, we write the sum of the age-structured mixing matrices as $\boldsymbol{\beta}(\phi)$.
To ensure that we can replicate the long-term dynamics of infection we allow the population to age. The aging process occurs annually (corresponding to the new school year in September) in which approximately one fifth of each age-group moves to the next oldest age cohort — small changes to the proportion moving between age-groups are made to keep the population size within each age-group constant.

**Capturing Quarantining**

One of the key characteristics of the COVID-19 pandemic in the UK has been the use of self-isolation and household quarantining to reduce transmission. We approximate this process by distinguishing between first infections (caused by infection related to any non-household mixing) and subsequent household infections (caused by infection due to household mixing). The first symptomatic case within a household (which might not be the first infection) has a probability (*H*) of leading to household quarantining; this curtails the non-household mixing of the individual and all subsequent infections generated by this individual.

In our notation, we let superscripts denote the first infection in a household (F), a subsequent infection from a symptomatic household member (SI) and a subsequent infection from an asymptomatic household member (SA); the first detected case in a household who is quarantined (QF) and all their subsequent household infections (QS). For a simple SEIR model (ignoring multiple E categories and age-structure) our extension would give:

$$
\begin{aligned}
\frac{dS}{dt} &= -(\lambda^F + \lambda^{SI} + \lambda^{SA} + \lambda^Q)S \\
\frac{dE^F}{dt} &= \lambda^F S - \alpha E^F \\
\frac{dE^{SI}}{dt} &= \lambda^{SI} S - \alpha E^{SI} \\
\frac{dE^{SA}}{dt} &= \lambda^{SA} S - \alpha E^{SA} \\
\frac{dE^Q}{dt} &= \lambda^Q S - \alpha E^Q \\
\frac{dI^F}{dt} &= d(1-H)\alpha E^F - \gamma I^F \\
\frac{dI^{SI}}{dt} &= d\alpha E^{SI} - \gamma I^{SI} \\
\frac{dI^{SA}}{dt} &= d(1-H)\alpha E^{SA} - \gamma I^{SA} \\
\frac{dI^{QF}}{dt} &= dH\alpha E^F - \gamma I^{QF} \\
\frac{dI^{QS}}{dt} &= d\alpha E^Q + dH\alpha E^{SA} - \gamma I^{QS} \\
\frac{dA^*}{dt} &= (1-d)\alpha E^* - \gamma A^* \\
\frac{dR}{dt} &= \gamma \sum_X I^X + A^X
\end{aligned}
$$

$$
\begin{aligned}
\text{where } \lambda^F &= (\beta^S + \beta^W + \beta^O)(I^F + I^{SI} + I^{SA} + \tau A^F + \tau I^{SI} + \tau A^{SA}) \\
\lambda^{SI} &= \beta^H I^F \\
\lambda^{SA} &= \beta^H A^F \\
\lambda^Q &= \beta^H A^{QF}
\end{aligned}
$$

This formation has been shown to be able to reduce *R* below one even when there is strong within household transmission, as infection from quarantined individuals cannot escape the household [3].

**Spatial Modelling**

Within England the model operates at the scale of NHS regions (East of England, London, Midlands, North East, North West, South East and South West). For simplicity and speed of simulation we assume that each of these regions acts independently and in isolation - we do not model the movement of people or infection across borders. In addition, the majority of parameters are regionally specific, reflecting different demographics, deprivation and social structures within each region. However, we include a hyper-prior on the shared parameters such that the behaviour of each region helps inform the value in others.

**Variant Modelling**

The model also captures the three main variants that have been responsible for most infections in England: the wildtype virus (encapsulating all pre-Alpha variants), the Alpha variant and the Delta variant. Each of these requires a replication of the infectious states for each variant type modelled. We assume that infection with each variant confers immunity to all variants, such that there is indirect competition for susceptible individuals. This competition is driven by the transmission advantage of each variant which is estimated by matching to the proportion of positive community PCR tests (Pillar

2 test) that are positive for the S-gene. The TaqPath system that is used for the majority of PCR tests in England is unable to detect the S-gene in Alpha variants, due to mutations in the S-gene. The switch from S-gene positive to S-gene negative and back to S-gene positive corresponds with the dominance of wildtype, Alpha and Delta variants. We infer the transmissibility of Alpha and Delta variants to be 52% (35-71%) and 156% (117-210%) greater than wildtype, respectively.

**Vaccination Modelling**

We capture vaccination using a leaky approach, although non-leaky (all-or-nothing) models produce extremely similar results over the time-scales considered. The model replicates the action of:

- first and second doses of vaccine, at rates $v_1$ and $v_2$ respectively which move susceptible individuals through to vaccinated states ($S_1$ and $S_2$) but have no impact on infected or recovered individuals;
- waning vaccine efficacy at rates $\omega_1$ and $\omega_2$, giving a two-step process from fully vaccinated to waned efficacy (in the equation below, for simplicity we assume everyone who gets a first dose of vaccine also gets a second, so that waning from state $S_1$ is unnecessary);
- waning immunity at rates $\Omega_1$ and $\Omega_2$ which are assumed to be slower than the waning of vaccine efficacy.

The model also needs to capture the total number of individuals who have been given a first or second dose of vaccine ($V_1$ or $V_2$ out of a total population size of $N$) to ensure that only individuals that have not been vaccinated are offered a first dose, and only individuals that have been vaccinated once are offered a second dose.

$$\begin{aligned}
\frac{dS}{dt} &= -v_1 \frac{S}{N - V_1} \\
\frac{dVS_1}{dt} &= v_1 \frac{S}{N - V_1} - v_2 \frac{VS_1}{V_1} \\
\frac{dVS_2}{dt} &= v_2 \frac{VS_1}{V_1} - \omega_1 VS_2 \\
\frac{dWS_1}{dt} &= \omega_1 VS_2 - \omega_2 WS_1 \\
\frac{dWS_2}{dt} &= \omega_2 WS_1
\end{aligned}$$

$$\begin{aligned}
\frac{dR}{dt} &= -\Omega_1 R \\
\frac{dWR_1}{dt} &= \Omega_1 R - \Omega_2 WR_1 \\
\frac{dWR_2}{dt} &= \Omega_2 WR_1
\end{aligned}$$

$$\begin{aligned}
\frac{dV_1}{dt} &= v_1 \\
\frac{dV_2}{dt} &= v_2
\end{aligned}$$

For those in the classes where the vaccines generate protection ($VS_1$, $VS_2$ and $WS_1$), the degree of protection is determined by the ratio of AstraZeneca (ChAdOx1) vaccine to mRNA vaccines (either Pfizer BNT162b2 or the Moderna COVID-19 vaccine) that has been given to that age group (see Table S1). If a vaccinated individual becomes infected, their probability of being admitted to hospital or dying - which normally only depends on age - is modified by the appropriate vaccine efficacy according to the ratio of the two vaccine types.

**Supplementary References**.


1. Moore S, Hill EM, Tildesley MJ, Dyson L, Keeling MJ. Vaccination and non-pharmaceutical interventions for COVID-19: a mathematical modelling study. Lancet Infect Dis. 2021;21: 793–802. doi:10.1016/S1473-3099(21)00143-2
2. Keeling MJ, Tildesley MJ, Atkins BD, Penman B, Southall E, Guyver-Fletcher G, et al. The impact of school reopening on the spread of COVID-19 in England. Philos Trans R Soc B Biol Sci. 2021;376: 20200261. doi:10.1098/rstb.2020.0261
3. Keeling MJ, Hill EM, Gorsich EE, Penman B, Guyver-Fletcher G, Holmes A, et al. Predictions of COVID-19 dynamics in the UK: Short-term forecasting and analysis of potential exit strategies. Flegg JA, editor. PLOS Comput Biol. 2021;17: e1008619. doi:10.1371/journal.pcbi.1008619
4. Dyson L, Hill EM, Moore S, Curran-Sebastian J, Tildesley MJ, Lythgoe KA, et al. Possible future waves of SARS-CoV-2 infection generated by variants of concern with a range of characteristics. Nat Commun. 2021;12: 5730. doi:10.1038/s41467-021-25915-7
5. Keeling MJ, Dyson L, Guyver-Fletcher G, Holmes A, Semple MG, Tildesley MJ, et al. Fitting to the UK COVID-19 outbreak, short-term forecasts and estimating the reproductive number. medRxiv. 2020; 2020.08.04.20163782. doi:10.1101/2020.08.04.20163782
6. Andrews N, Tessier E, Stowe J, Gower C, Kirsebom F, Simmons R, et al. Vaccine effectiveness and duration of protection of Comirnaty, Vaxzevria and Spikevax against mild and severe COVID-19 in the UK. medRxiv. 2021; 2021.09.15.21263583. doi:10.1101/2021.09.15.21263583
7. Pouwels KB, Pritchard E, Matthews PC, Stoesser N, Eyre DW, Vihta K-D, et al. Effect of Delta variant on viral burden and vaccine effectiveness against new SARS-CoV-2 infections in the UK. Nat Med. 2021. doi:10.1038/s41591-021-01548-7
8. Lin D-Y, Gu Y, Wheeler B, Young H, Holloway S, Khan S, et al. Effectiveness of Covid-19 Vaccines in the United States Over 9 Months: Surveillance Data from the State of North Carolina. medRxiv. 2021 2021.10.25.21265304. doi: 10.1101/2021.10.25.21265304
9. Baguelin M, Camacho A, Flasche S, Edmunds WJ. Extending the elderly- and risk-group programme of vaccination against seasonal influenza in England and Wales: a cost-effectiveness study. BMC Med. 2015;13: 236. doi:10.1186/s12916-015-0452-y
10. Hill EM, Petrou S, Forster H, de Lusignan S, Yonova I, Keeling MJ. Optimising age coverage of seasonal influenza vaccination in England: A mathematical and health economic evaluation. Althouse BM, editor. PLOS Comput Biol. 2020;16: e1008278. doi:10.1371/journal.pcbi.1008278
11. National Institute for Health and Care Excellence. Guide to the processes of technology appraisal. 2018. Available: https://www.nice.org.uk/Media/Default/About/what-we-do/NICE-guidance/NICE-technology-appraisals/technology-appraisal-processes-guide-apr-2018.pdf
12. Sandmann FG, Davies NG, Vassall A, Edmunds WJ, Jit M, Sun FY, et al. The potential health and economic value of SARS-CoV-2 vaccination alongside physical distancing in the UK: a transmission model-based future scenario analysis and economic evaluation. Lancet Infect Dis. 2021;21: 962–974. doi:10.1016/S1473-3099(21)00079-7
13. Torres-Rueda S, Sweeney S, Bozzani F, Vassall A. The health sector cost of different policy responses to COVID-19 in low- and middle-income countries. medRxiv. 2020; 2020.08.23.20180299. doi:10.1101/2020.08.23.20180299
14. Prem K, Cook AR, Jit M. Projecting social contact matrices in 152 countries using contact surveys and demographic data. Halloran B, editor. PLOS Comput Biol. 2017;13: e1005697. doi:10.1371/journal.pcbi.1005697
15. Mossong J, Hens N, Jit M, Beutels P, Auranen K, Mikolajczyk R, et al. Social Contacts and Mixing Patterns Relevant to the Spread of Infectious Diseases. Riley S, editor. PLoS Med. 2008;5: e74. doi:10.1371/journal.pmed.0050074
